# Assessing the Feasibility of Large Language Models to Identify Top Research Priorities in Enhanced External Counterpulsation

**DOI:** 10.1101/2024.06.01.24308314

**Authors:** Shengkun Gai, Fangwan Huang, Xuanyun Liu, Ryan G. Benton, Glen M. Borchert, Jingshan Huang, Xiuyu Leng

**Affiliations:** Department of Cardiology, The First Affiliated Hospital, Sun Yat-sen University, Guangzhou, China; Department of Cardiology, Linfen People’s Hospital, Shanxi, China; College of Computer and Data Science, Fuzhou University, Fuzhou, China; School of Computing, University of South Alabama, Mobile, AL, U.S.A.; College of Medicine, University of South Alabama, Mobile, AL, U.S.A.; School of Computing and College of Medicine, University of South Alabama, Mobile, AL, U.S.A.

**Keywords:** EECP, Research Priorities, Large Language Models, ChatGPT, Ernie Bot, Artificial Intelligence

## Abstract

Enhanced External Counterpulsation (EECP), as a non-invasive, cost-effective, and efficient adjunctive circulatory technique, has found increasingly widespread applications in the cardiovascular field. Numerous basic research and clinical observations have extensively confirmed the significant advantages of EECP in promoting blood flow perfusion to vital organs such as the heart, brain, and kidneys. However, many potential mechanisms of EECP remain insufficiently validated, necessitating researchers to dedicate substantial time and effort to in-depth investigations. In this work, we attempted to use large language models (such as ChatGPT and Ernie Bot) to identify top research priorities in five key topics in the field of EECP: mechanisms, device improvements, cardiovascular applications, neurological applications, and other applications. After generating specific research priorities in each domain through language models, we invited a panel of nine experienced EECP experts to independently evaluate and score them based on four parameters: relevance, originality, clarity, and specificity. Notably, average and median scores for these evaluation parameters were high, indicating a strong endorsement from experts in the EECP field. Although further validation and refinement are required, this study preliminarily suggests that large language models like ChatGPT and Ernie Bot could serve as powerful tools for identifying and prioritizing research priorities in the EECP domain.

## 1 INTRODUCTION

Enhanced External Counterpulsation (EECP) is a non-invasive adjunctive circulatory technique that inflates and deflates cuffs wrapped around the limbs and buttocks in sync with the cardiac cycle under electrocardiographic gating control. EECP has been shown clinically to significantly improve organ perfusion, regulate endothelial function, combat coronary artery atherosclerosis, treat complications of diabetes and sudden sensorineural hearing loss, among other benefits, through a series of mechanisms [1-3]. While evidence suggests there is still a great deal of untapped potential for external counterpulsation, traditional approaches to identifying research priorities for EECP rely mainly on expert opinion and consensus building which are often labor-intensive and biased. In recent years, natural language processing (NLP) technology [4] has been increasingly recognized as a new means of identifying research priorities. Large language models (LLMs) such as ChatGPT [6] and Ernie Bot [7], which are trained on extensive text data, possess the ability to understand human-like language and have demonstrated significant potential in proposing and prioritizing research priorities [5]. In this work, ChatGPT and Ernie Bot were evaluated for their effectiveness in identifying primary research priorities related to EECP technology. Five key areas were examined: mechanisms, device enhancements, cardiovascular applications, neurological applications, and other applications. Utilizing ChatGPT and Ernie Bot, specific research priorities in these domains were generated after which experienced EECP experts reviewed and then rated them to assess their relevance and importance.

## 2 RELATED WORK

Large language models have shown broad applicability in entertainment, education, and customer service, but their potential in the medical field remains largely untapped. Given the high standards for information quality and communication reliability in medicine, the application of large language models requires careful consideration. In recent years, scholars have begun to explore the use of large language models in medicine, yielding promising results. In the field of cardiology, Gala et al. [8] believe that LLMs can analyze a large number of research papers and medical record resources to help clinicians keep up with the latest advances in cardiology. But they also point to the limitations of LLMs in explaining cultural or emotional factors that may influence medical practice. Cascella et al. [9] explored ChatGPT’s reasoning abilities on public health topics. Through a question-and-answer session, ChatGPT listed four possible research topics. While some of ChatGPT’s responses may be stereotyped and depend on the prompts, it can be used to summarize the scientific literature and generate new research hypotheses. Additionally, George et al. [10] proposed that large language models can serve as a supplementary resource to traditional medical tools, improving the efficiency and productivity of medical practices. Unfortunately, these studies do not provide a quantitative assessment of the LLMs’ ability to identify medical research priorities.

Importantly, in order to assess the effectiveness of LLMs in the medical domain, it is essential to conduct statistical analyses on numerical results obtained from experiments and/or surveys. In evaluating the pertinent literature on LLMs, Tang et al. [11] invited field experts to assess the summary quality of LLMs by using a five-point Likert scale along four dimensions: coherence, factual consistency, comprehensiveness, and harmfulness. The Man-Whitney U test was used to assess the differences in response between GPT-3.5 and ChatGPT. Michael et al. [12] employed average scoring and fixed-effects consistency to calculate the Intraclass Correlation Coefficient (ICC), investigating the potential application of artificial intelligence-based LLMs in the realm of medical ethics. Similarly, Dave et al. [13] utilized Pearson and Spearman coefficients to juxtapose the assessment outcomes of large language models against the evaluations of medical professionals, thereby further substantiating their dependability. Furthermore, besides correlation analysis, similarity metrics are frequently utilized to gauge the efficacy of LLMs. For example, in 2024, Sebastian et al. [14] evaluated the pairwise accuracy between LLMs and human assessments by analyzing the cosine similarity matrix. In measuring factual knowledge within LLMs, Pezeshkpour [15] successfully utilized Kullback-Leibler (KL) divergence to analyze the predictive probability distributions of the model before and after instilling target knowledge. Guo et al. [16], in investigating bias issues within large pre-trained language models, used the Jensen-Shannon (JS) divergence to measure the consistency between different demographic distributions, offering a robust tool for reducing human-like biases and unwanted societal stereotypes. JS divergence is an improved version of KL divergence, being symmetric whereas KL divergence is asymmetric, rendering JS divergence more precise in discerning similarity.

## 3 METHODS

### Research priorities

We leveraged ChatGPT (based on GPT-3.5)which has captured 100 million users worldwide and Ernie Bot 3.5 which more Popular in China to generate research priorities in five key topics (Table 1 and Table 2, respectively) pertaining to EECP mechanisms [1], [17], structural enhancements, applications in cardiovascular domains [3], [18], [19], neurological applications [20], [21], and other applications [3], [22], [23].

**Table 1.**
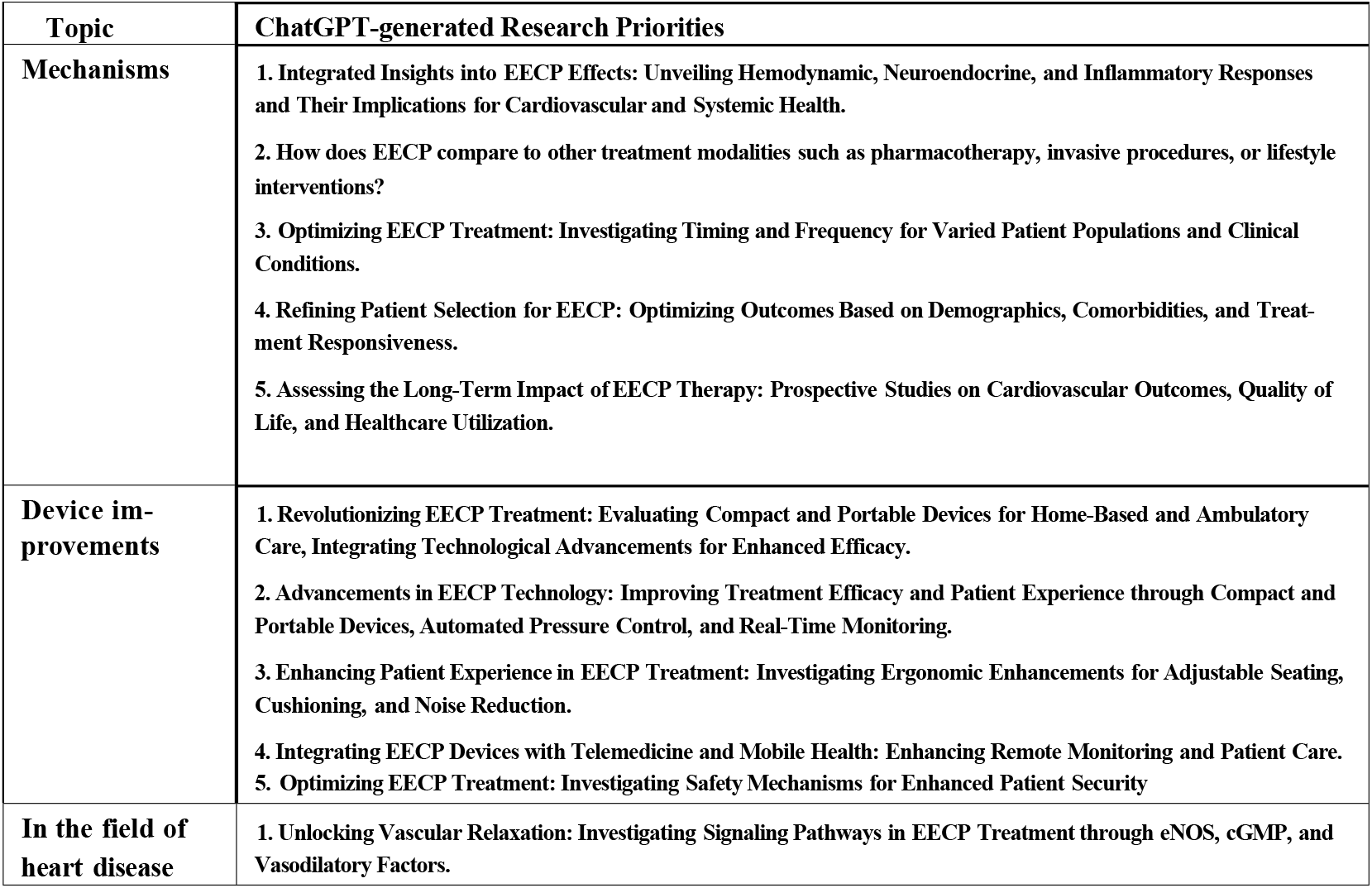
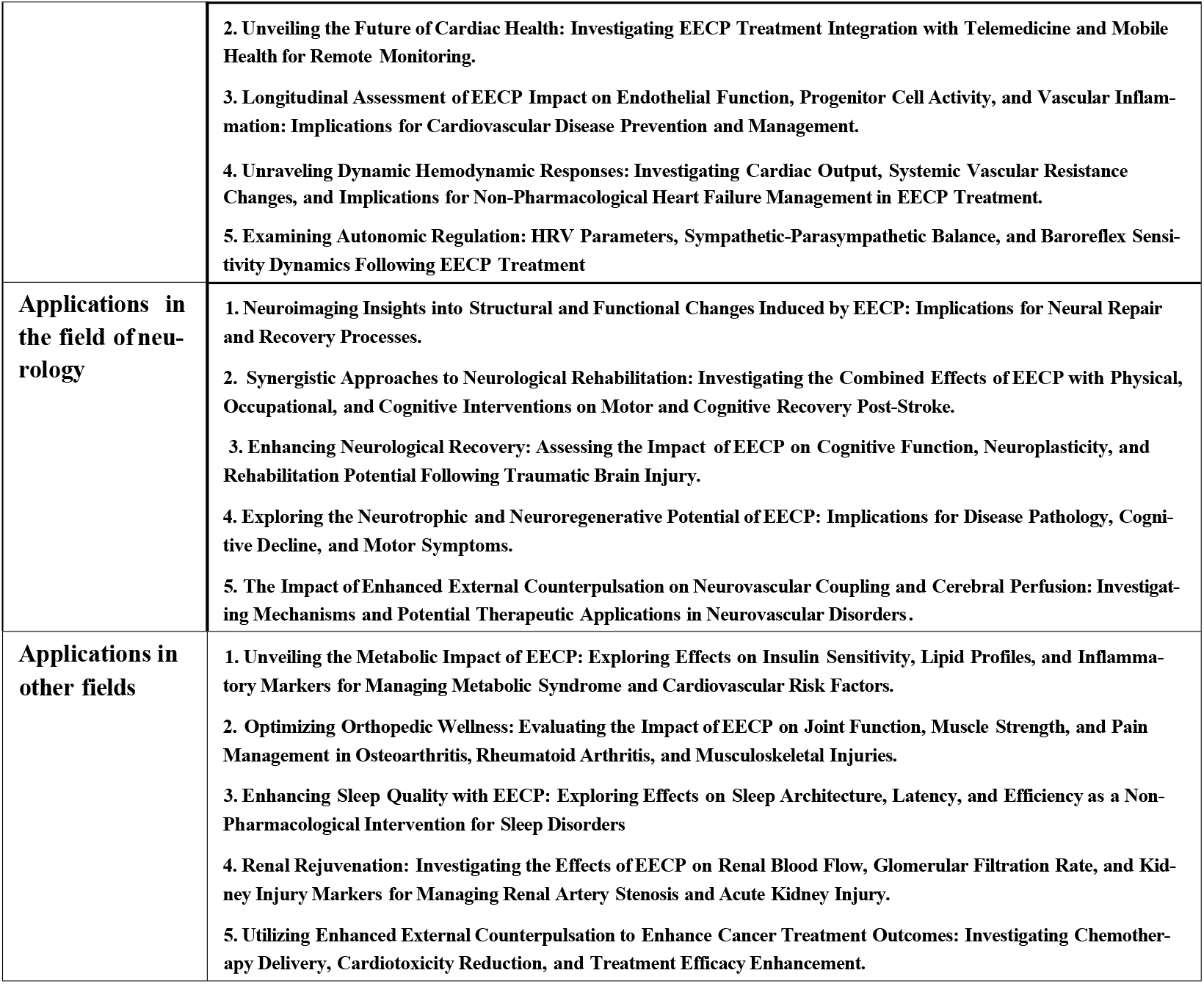
ChatGPT-generated research prioritieson five key topics in the field of EECP research

**Table 2.**
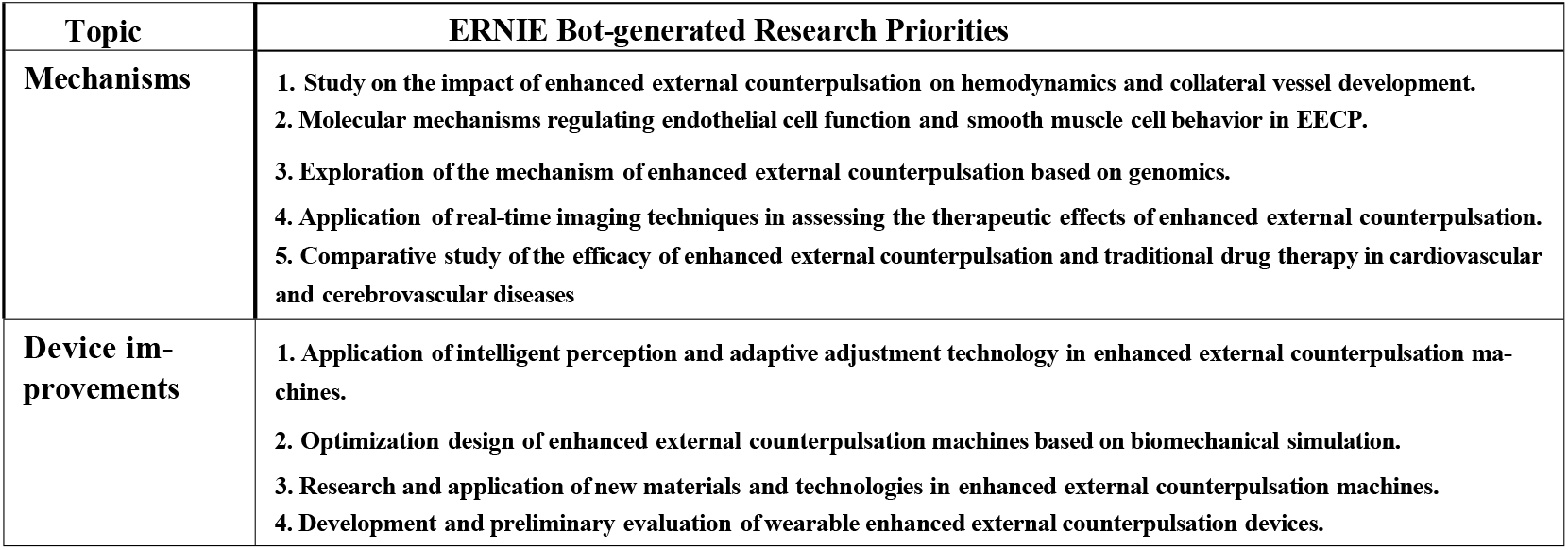
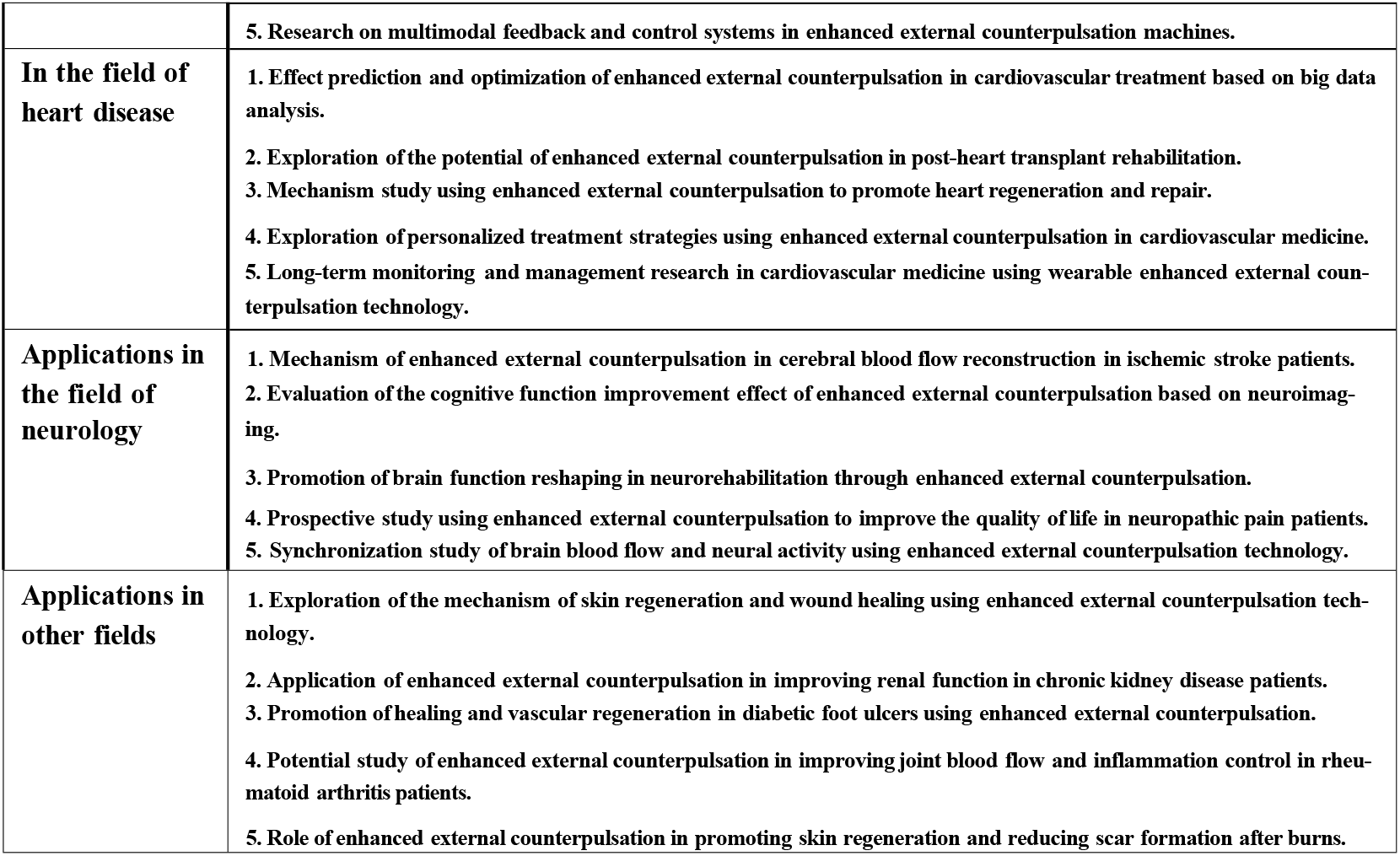
ERNIE bot-generated research priorities on five key topics in the field of EECP research

### Expert evaluation

The expert evaluation panel was comprised of nine highly experienced EECP specialists as evidenced by panelists having authored an average of twenty relevant research publications in the field. Panelists reviewed and assessed the research inquiries presented by ChatGPT and Ernie Bot independently. Experts rated fvie priorities on four parameters (relevance, originality, clarity, and specificity) using a 1-5 scale with 5 representing the highest score. ChatGPT and Ernie Bot-generated priorities were then compared to current EECP research queries identified through manual literature review. Importantly, in order to ensure the objectivity and relevance of responses, ChatGPT and Ernie Bot were instructed to treat each key topic as an independent query, thereby eliminating potential biases that may have existed in previous conversations.

## 4 STATISTICAL ANALYSIS

Standard statistical methods were utilized for both data collection and analysis with all statistical analyses carried out using IBM SPSS Statistics version 25 and Python 3.10. Initially, descriptive statistical methods were employed to provide a summary of the data, including measures such as mean, standard deviation (SD), and median. Following this, the study utilized “divergence” to assess the similarity between ratings provided by experts in EECP and queries generated by two large language models. In the realm of data mining, JS divergence was computed to evaluate the similarity of ratings among evaluators using a rating table structured with evaluators as column attributes. JS divergence values from 0 to 1, with smaller values indicating greater similarity between ratings. Additionally, Spearman’s rank correlation coefficient and Kendall’s τ coefficient were also used to evaluate pairwise correlations between parameters. Positive coefficients indicate a positive correlation, while negative coefficients imply a negative correlation. The closer the coefficient is to 1 the stronger the correlation.

## 5 RESULTS

The statistical analysis shows high reliability for the questionnaires assessing ChatGPT and Ernie Bot, with Cronbach’s alpha coefficients of 0.978 and 0.971, respectively. Both coefficients exceed the 0.8 threshold, indicating strong survey reliability. This suggests that the questionnaires effectively reflect the proficiency of ChatGPT and Ernie Botin determining research priorities for EECP.

Based on this, the study conducted data analysis on the ratings provided by the 9 evaluators from three perspectives: (1) descriptive statistics; (2) similarity of ratings among evaluators; and (3) rank correlation of evaluation metrics. The data analysis tools utilized were IBM SPSS Statistics Version 25 and Python 3.10.

### 5.1 Descriptive statistics

In-depth descriptive statistical analyses of evaluation metrics are presented in Tables 3 to 5. The major models performed best in relevance, with originality close behind. While originality exhibited the largest standard deviation, suggesting significant variation in expert opinions regarding originality, clarity demonstrated the smallest standard deviation, indicating minimal fluctuations in scores for each question. Additionally, variations in performance between the two models (ChatGPT and Ernie Bot) across different evaluation metrics and topics can be observed. Concerning relevance, Ernie Bot’s average score slightly exceeds ChatGPT’s, suggesting a slight advantage in addressing user-related questions, although this was not statistically significant. In terms of originality, ChatGPT’s score was slightly less than Ernie Bot’s, with a higher fluctuation in scoring standard deviation, indicating some disagreement among experts regarding the originality of ChatGPT’s queries. Both models demonstrate similar performance in clarity and specificity, indicating their similarity in providing clear and specific answers. Results of scores from EECP experts for all priorities are visually presented in Figure 1 with the outermost rings corresponding to the highest score of 5 and inner rings indicating lower scores.

**Table 3.**
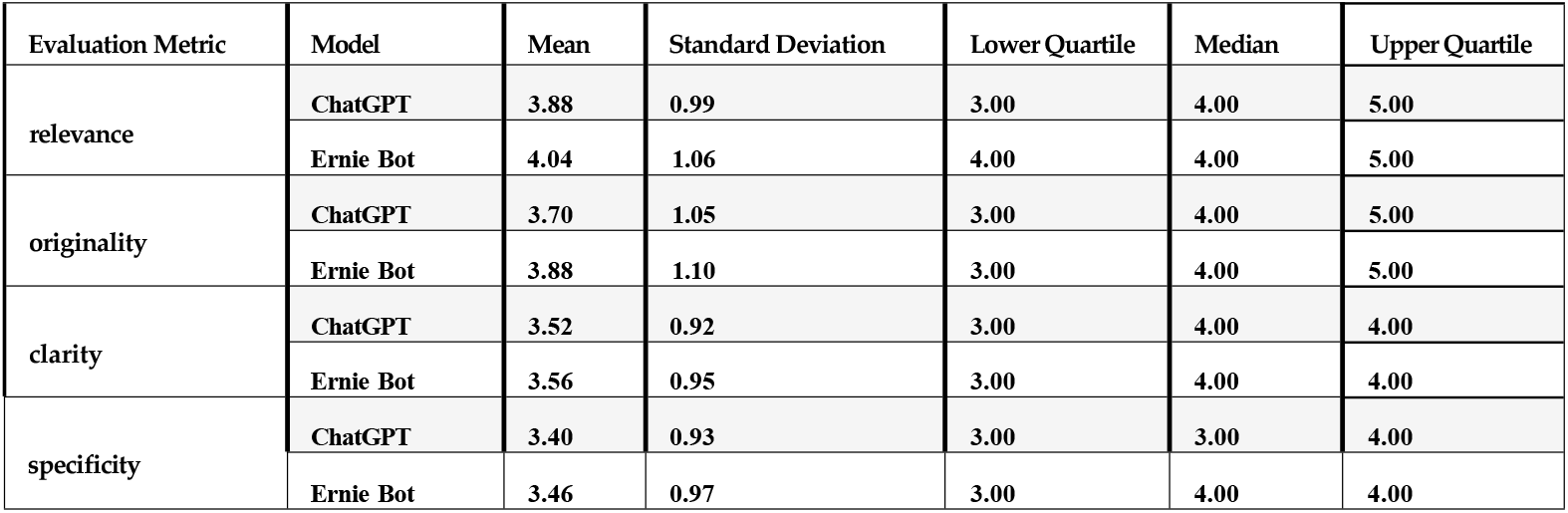
Descriptive statistics of evaluation metrics

**Figure 1.**
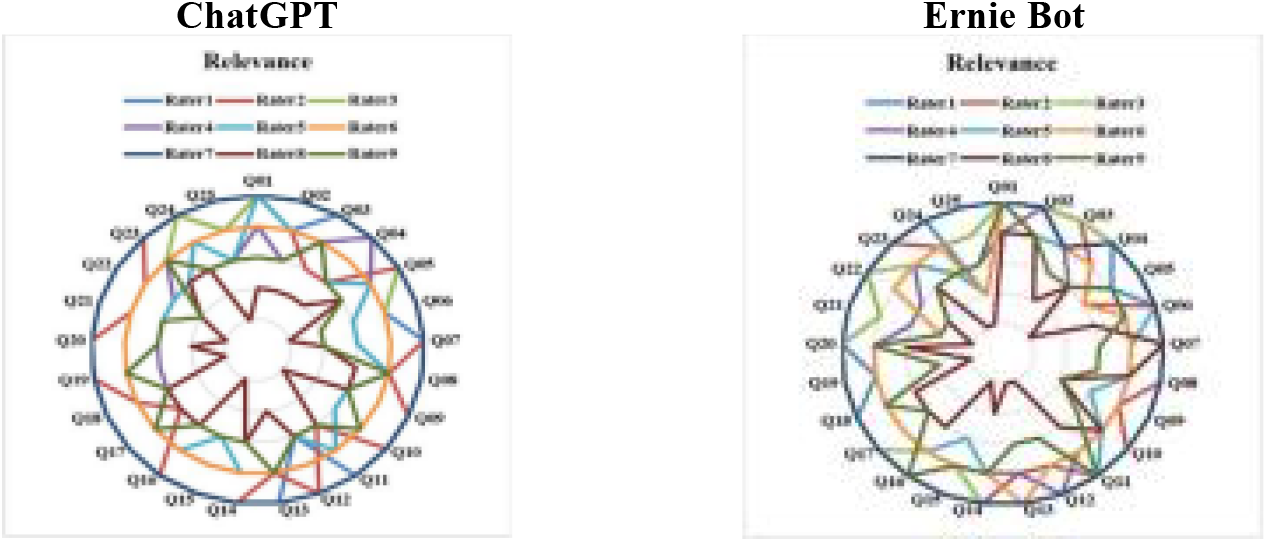
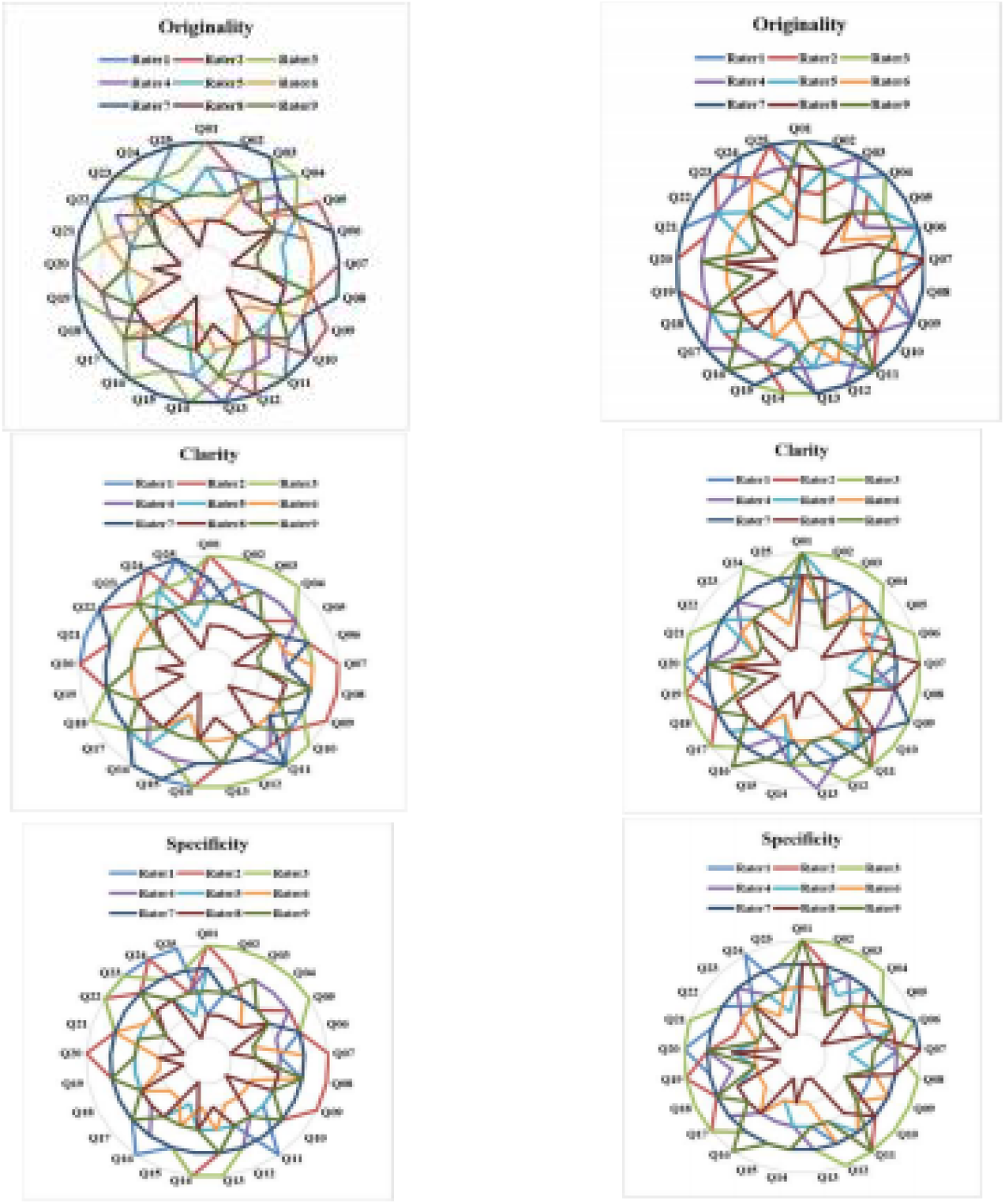
Ratings of 25 research focal points by nine evaluators based on four criteria.

Table 4 presents the scores given by different raters for the ChatGPT and Ernie Bot models. The analysis shows that in the evaluations of most raters, ChatGPT and Ernie Bot have similar average scores indicating a certain level of competitiveness in overall performance. However, it is worth noting that in the ratings ofRater3 and Rater4, Ernie Bot’s average score was significantly higher than ChatGPT’s, reflecting a more outstanding performance of Ernie Bot from the perspectives of these two raters. In terms of score stability, there were differences between the two models among different raters. Specifically, in the evaluations of Rater3 and Rater4, Ernie Bot had a lower standard deviation, indicating more stable scores and consistent performance. Conversely, Rater8’s Ernie Bot scores demonstrated significantly higher standard deviation. In contrast, ChatGPT’s standard deviation among multiple raters was relatively more consistent, although overall score stability was slightly inferior to Ernie Bot’s performance for a subset of raters. These differences in evaluation may stem from personal preferences, evaluation criteria, and model performance across different topics.

**Table 4.** Descriptive statistics of evaluator

| Evaluator | Model | Mean | Standard Deviation | Lower Quartile | Median | Upper Quartile |
| --- | --- | --- | --- | --- | --- | --- |
| Rater1 | ChatGPT | 4.34 | 0.71 | 4.00 | 4.00 | 5.00 |
|  | Ernie Bot | 4.10 | 0.78 | 4.00 | 4.00 | 5.00 |
| Rater2 | ChatGPT | 4.30 | 0.70 | 4.00 | 4.00 | 5.00 |
|  | Ernie Bot | 4.20 | 0.67 | 4.00 | 4.00 | 5.00 |
| Rater3 | ChatGPT | 4.37 | 0.49 | 4.00 | 4.00 | 5.00 |
|  | Ernie Bot | 4.75 | 0.44 | 4.25 | 5.00 | 5.00 |
| Rater4 | ChatGPT | 3.50 | 0.54 | 3.00 | 3.00 | 4.00 |
|  | Ernie Bot | 3.81 | 0.61 | 3.00 | 4.00 | 4.00 |
| Rater5 | ChatGPT | 3.16 | 0.58 | 3.00 | 3.00 | 3.00 |
|  | Ernie Bot | 3.37 | 0.86 | 3.00 | 3.00 | 4.00 |
| Rater6 | ChatGPT | 3.17 | 0.77 | 3.00 | 3.00 | 4.00 |
|  | Ernie Bot | 3.14 | 0.79 | 3.00 | 3.00 | 4.00 |
| Rater7 | ChatGPT | 4.44 | 0.67 | 4.00 | 5.00 | 5.00 |
|  | Ernie Bot | 4.50 | 0.52 | 4.00 | 5.00 | 5.00 |
| Rater8 | ChatGPT | 2.20 | 0.80 | 2.00 | 2.00 | 3.00 |
|  | Ernie Bot | 2.44 | 1.21 | 1.00 | 3.00 | 3.00 |
| Rater9 | ChatGPT | 3.16 | 0.61 | 3.00 | 3.00 | 4.00 |
|  | Ernie Bot | 3.31 | 0.92 | 3.00 | 3.00 | 4.00 |

In all topics (refer to Table 5), Ernie Bot consistently received higher average scores than ChatGPT, suggesting a relative advantage in overall performance. Although their performances in terms of median scores were similar, Ernie Bot achieved an upper quartile score of 5.00 in specific topics such as mechanisms, device improvements and applications in neurology, indicating higher recognition in these areas. Meanwhile ChatGPT’s standard deviation across multiple topics was slightly lower than Ernie Bot’s, suggesting relatively better score stability. However, this difference was not significant. Notably, clear domain-specific differences were observed, while Ernie Bot’s average score significantly surpassed ChatGPT’s in structural improvements and applications in neurology domains, ChatGPT demonstrated superior performance in other domains.

**Table 5.** Descriptive statistics of topic

| topic | Model | Mean | Standard Deviation | Lower Quartile | Median | Upper Quartile |
| --- | --- | --- | --- | --- | --- | --- |
| mechanisms | ChatGPT | 3.63 | 0.95 | 3.00 | 4.00 | 4.00 |
|  | Ernie Bot | 3.83 | 0.98 | 3.00 | 4.00 | 5.00 |
| device improvements | ChatGPT | 3.52 | 1.03 | 3.00 | 4.00 | 4.00 |
|  | Ernie Bot | 3.94 | 0.90 | 3.00 | 4.00 | 5.00 |
| in the field of heart disease | ChatGPT | 3.69 | 0.98 | 3.00 | 4.00 | 4.00 |
|  | Ernie Bot | 3.68 | 1.12 | 3.00 | 4.00 | 4.75 |
| in the field of neurology | ChatGPT | 3.67 | 0.91 | 3.00 | 4.00 | 4.00 |
|  | Ernie Bot | 3.85 | 0.99 | 3.00 | 4.00 | 5.00 |
| the other field | ChatGPT | 3.62 | 1.06 | 3.00 | 4.00 | 4.00 |
|  | Ernie Bot | 3.38 | 1.14 | 3.00 | 3.00 | 4.00 |

### 5.2 Similarity of raters’ scores

Regarding the similarity of raters’ scores, we calculated the JS divergence of scores between each pair of raters for ChatGPT and Ernie Bot (see Figure 2 for details). The results indicate that the JS divergence range of scores for ChatGPT is [0, 0.102], while for Ernie Bot, it is [0, 0.148]. Since a smaller JS divergence value indicates higher similarity, it can be concluded that the evaluations of these two large language models by raters exhibit relatively high consistency. It is worth noting that, for both ChatGPT and Ernie Bot, the similarity of scores between rater 8 and other raters is the lowest. From Figure 1, it is evident that the scores given by rater 8 are significantly lower than those given by other raters. Further analysis of the data in Table 4 reveals that the average scores given by rater 8 for both ChatGPT and Ernie Bot are the lowest (2.20 and 2.44 respectively), and they have the highest standard deviations (0.80 and 1.21 respectively). Excluding the influence of rater 8’s scores, the upper limit of the JS divergence of scores for ChatGPT would decrease from 0.102 to 0.052, and from 0.148 to 0.063 for Ernie Bot.

**Figure 2.**
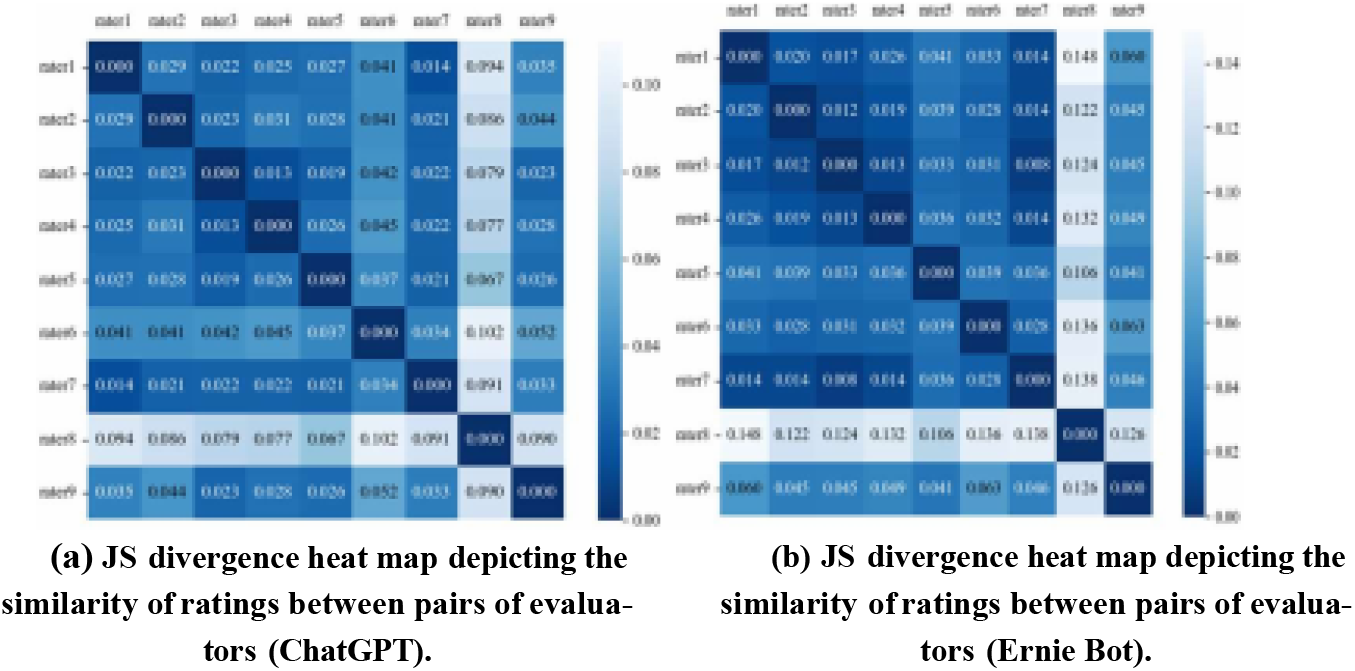
JS divergence heat map depicting the similarity of ratings between pairs of evaluators.

### 5.3 Correlation of evaluation metrics

In terms of the correlation of evaluation metrics, we calculated both the Spearman [24] and Kendall [25] coefficients between pairs of evaluation metrics in the scoring results for ChatGPT and Ernie Bot (see Tables 6 and 7). These analyses passed significance tests, with all p-values below 0.01 indicating a significant positive correlation between relevance, originality, clarity, and specificity. This implies that when evaluating these two models, the score trends among these metrics were consistent, demonstrating high consistency and reliability. That said, ChatGPT exhibited a lower correlation between originality and relevance, while Ernie Bot showed a lower correlation in the analysis of specificity and relevance. The clarity of both models was highly correlated with relevance and/or specificity.

**Table 6.** Rank correlation coefficients between evaluation metrics (ChatGPT)

| Spearman's coefficient | relevance | originality | clarity | specificity |
| --- | --- | --- | --- | --- |
| relevance | 1 | 0.778** | 0.722** | 0.670** |
| originality | 0.778** | 1 | 0.780** | 0.772** |
| clarity | 0.722** | 0.780** | 1 | 0.883** |
| specificity | 0.670** | 0.772** | 0.883** | 1 |
| Kendall's coefficient | relevance | originality | clarity | specificity |
| relevance | 1 | 0.713** | 0.667** | 0.605** |
| originality | 0.713** | 1 | 0.726** | 0.711** |
| clarity | 0.667** | 0.726** | 1 | 0.840** |
| specificity | 0.605** | 0.711** | 0.840** | 1 |

Note: \*\* Significance at the 0.01 level (two-tailed).

**Table 7.** Rank correlation coefficients between evaluation metrics (Ernie Bot)

| Spearman's coefficient | relevance | originality | clarity | specificity |
| --- | --- | --- | --- | --- |
| relevance | 1 | 0.692** | 0.695** | 0.708** |
| originality | 0.692** | 1 | 0.740** | 0.769** |
| clarity | 0.695** | 0.740** | 1 | 0.876** |
| specificity | 0.708** | 0.769** | 0.876** | 1 |
| Kendall's coefficient | relevance | originality | clarity | specificity |
| relevance | 1 | 0.643** | 0.646** | 0.650** |
| originality | 0.643** | 1 | 0.686** | 0.707** |
| clarity | 0.646** | 0.686** | 1 | 0.846** |
| specificity | 0.650** | 0.707** | 0.846** | 1 |

Note: \*\* Significance at the 0.01 level (two-tailed).

## 6 DISCUSSION

This work assessed the ability of ChatGPT and Ernie Bot to generate research priorities in the field of EECP, covering mechanisms, structural improvements, applications in cardiology, applications in neurology, and applications in other fields. Both models demonstrated significant potential in consistently generating relevant and clear research priorities , which could offer valuable new tools for EECP research. That said, both scored relatively low in specificity, possibly due to limitations in handling domain-specific knowledge, indicating a need for improvement in accuracy and precision. To enhance their performance, fine-tuning with domain-specific data and expert knowledge will likely be required. While both models lacked originality in their responses, relying heavily on learned information and language patterns, future research should focus on enhancing their creativity to generate more unique research questions in the EECP field.

Notably, this study also compared the performances of Ernie Bot and ChatGPT, two prominent language systems. Ernie Bot demonstrated a slight but definitive advantage in terms of relevance, possibly due to its more precise semantic understanding and higher matching with user needs. In terms of originality, ChatGPT scored slightly lower with more fluctuation, indicating some disagreement among evaluators regarding its ability to offer novel and unique perspectives. This variance might stem from differences in the models’ performance across different contexts or from evaluators’ subjective criteria, such as their acceptance of research priorities that challenge existing cognitive frameworks or their willingness to explore unknown areas of study. In contrast, Ernie Bot received more consistent recognition for its originality, likely due to its more flexible and innovative thinking patterns. Regarding clarity and specificity, both models performed equally well, demonstrating high levels of proficiency. This suggests that they excel in providing clear, understandable responses and specific, detailed explanations, which are equally important for large language models as users often expect answers that are both clear and specific to better understand and apply the provided information.

From the evaluators’ perspective, most evaluators held similar views on the performance of the two models. However, in certain specific cases, such as Rater3 and Rater4, Ernie Bot received higher scores. Additionally, as compared to other raters, Rater8’s scores were significantly lower and deviated more substantially, and exclusion of Rater8 increased the performance of both models.

In certain specific topics such as mechanisms, applications in neurology, and cardiovascular applications, Ernie Bot performed better whereas ChatGPT’s performance slightly surpassed that of Ernie Bot in others, indicating that each model has its strengths and weaknesses in different domains and application scenarios. Therefore, future research could further explore how to integrate the strengths of both models to enhance the performance and effectiveness of large language models in practical applications.

While the study yields promising outcomes, there were some clear limitations. Firstly, the expert panels involved may not fully represent the broader research community which could have influenced evaluation outcomes. Secondly, the use of subjective ratings could have introduced potential bias and variability in assessing ChatGPT and Ernie Bot’s performance. Lastly of note, the models may lack access to the latest biomedical literature, impacting question generation. If true, integrating domain-specific APIs with updated information could enhance research quality.

## 7 CONCLUSION

Overall, this assessment of ChatGPT and Ernie Bot as generators of research priorities for Enhanced External Counterpulsation (mechanisms, device improvements, applications in cardiovascular medicine, applications in neurology, and applications in other non-cardiovascular and non-neurological fields) produced some promising results. Both models have demonstrated the capacity to generate high-quality research priorities in these areas, indicating their potential value as tools to drive research not only in EECP but also in broader medical fields through streamlining the process of identifying crucial research priorities and thereby save considerable time and effort. While there is room for improvement in terms of specificity and originality, both models have shown a capability to produce diverse, relevant, and coherent research priorities, likely aiding advancements in EECP research. Each model has its strengths in various domains and application scenarios, and further exploration could focus on leveraging these strengths to enhance the overall effectiveness of large language models in practical settings. In conclusion, our findings suggest that ChatGPT and Ernie Bot are poised to become valuable assistants for researchers in the EECP field and likely other medical domains, offering new momentum for scientific progress.

## Data Availability

All relevant data are within the manuscript and its Supporting Information files.

## Notes

### Competing Interest Statement

The authors have declared no competing interest.

### Funding Statement

The author(s) received no specific funding for this work.

### Author Declarations

This experiment does not require ethical approval

